# Suboptimal Sleep Duration is Associated with Poorer Neuroimaging Brain Health Profiles

**DOI:** 10.1101/2023.04.20.23288891

**Authors:** Santiago Clocchiatti-Tuozzo, Cyprien Rivier, Daniela Renedo, Victor M Torres Lopez, Jacqueline Geer, Brienne Miner, Henry Yaggi, Adam de Havenon, Sam Payabvash, Kevin N Sheth, Thomas M Gill, Guido J Falcone

## Abstract

**Background:** Cardiovascular health optimization during middle age benefits brain health. The American Heart Association’s Life’s Simple 7 recently added sleep duration as a key determinant of cardiovascular health becoming the Life’s Essential 8. We tested the hypothesis that suboptimal sleep duration is associated with poorer neuroimaging brain health profiles in asymptomatic middle-aged adults.

**Methods:** We conducted a prospective MRI neuroimaging study in middle-aged persons without stroke, dementia, or multiple sclerosis enrolled in the UK Biobank. Self-reported sleep duration was categorized as short (<7 hours), optimal (7-<9 hours), or long (≥9 hours). Evaluated neuroimaging markers of brain health included white matter hyperintensities (presence and volume) and diffusion tensor imaging metrics (fractional anisotropy and mean diffusivity) evaluated in 48 distinct neuroanatomical regions. We used multivariable logistic and linear regression models, as appropriate, to test for association between sleep duration and neuroimaging markers of brain health.

**Results:** We evaluated 39,502 middle-aged persons (mean age 55, 53% female). Of these, 28,712 (72.7%) had optimal, 8,422 (21.3%) short, and 2,368 (6%) long sleep. Compared to optimal sleep, short sleep was associated with higher risk (OR 1.11; 95% CI 1.05-1.17; P<0.001) and larger volume (beta=0.06, SE=0.01; P<0.001) of white matter hyperintensities, while long sleep was associated with higher volume (beta=0.04, SE=0.02; P=0.01) but not higher risk (P>0.05) of white matter hyperintensities. Short (beta=0.03, SE=0.01; P=0.004) and long sleep (beta=0.07, SE=0.02; P<0.001) were associated with worse fractional anisotropy, while only long sleep associated with worse mean diffusivity (beta=0.05, SE=0.02; P=0.005).

**Conclusions:** Among middle-aged adults without clinically observed neurological disease, suboptimal sleep duration is associated with poorer neuroimaging brain health profiles. Because the evaluated neuroimaging markers precede stroke and dementia by several years, our findings support early interventions aimed at correcting this modifiable risk factor.

## Introduction

Sleep is a vital neurophysiological process linked to various health outcomes. Evidence shows that both short and long sleep durations are associated with higher risks, including all-cause mortality, coronary heart disease, and stroke.^1^ The American Heart Association recently introduced the Life’s Essential 8,^2^ a construct derived from the Life’s Simple 7 that highlights critical determinants of cardiovascular health. Importantly, it added sleep duration as a new factor. Recent research indicates that optimizing cardiovascular health in middle age leads to significant brain health benefits later in life.^3^ Our hypothesis is that suboptimal sleep duration (short and long) results in poorer neuroimaging brain health profiles among middle-aged individuals without a diagnosis of stroke, dementia, or multiple sclerosis.

## Methods

### Study design

We conducted a nested, prospective neuroimaging study within the UK Biobank (UKB), a large population-based cohort study in the United Kingdom.^4^ Between March 2006 and October 2010, the UKB enrolled 502,480 community-dwelling persons aged 40 to 69 years. After enrollment, a random subset of these participants was invited to participate in a neuroimaging study and completed a brain magnetic resonance imaging (MRI). Given that our primary focus was persons without clinically evident neurological disease, we excluded those who had a history of stroke, dementia, or multiple sclerosis.

### Exposure ascertainment and modeling

During the baseline interview, UKB participants were asked about the average number of hours of total sleep per day, including daytime napping.^4^ Following the classification scheme proposed by the American Heart Association,^2^ we categorized sleep duration as short (<7 hours), optimal (7 to <9 hours) and long (>=9 hours) sleep.

### Neuroimaging measurements

A detailed description of the MRI acquisition protocol is available elsewhere.^5^ Briefly, brain MRIs were completed using a Siemens Skyra 3T. The UKB research team centrally segmented the radiological phenotypes evaluated here:^6,7^ white matter hyperintensities (WMH)^8-10^ and white matter disintegrity, the latter evaluated via the diffusion tensor imaging metrics fractional anisotropy (FA) and mean diffusivity (MD) evaluated at 48 specific neuroanatomical areas.^9,11^ We used a previously validated method that calculates the first principal component of FA/MD measurements across all evaluated neuroanatomical regions.^12^ Within this framework, higher FA and MD scores reflect higher white matter disintegrity.

### Covariates

To reflect social deprivation, we used the Townsend Deprivation Index,^13^ a validated measure of material deprivation that uses 4 variables: unemployment, non-car ownership, non-home ownership, and household overcrowding.

### Statistical analyses

For WMH volume, we examined two outcomes: none versus any (dichotomous) and overall burden (continuous). We defined any WMH as having >5 cubic centimeters (CC) of WMH to minimize false positives. We initially used Chi-square and ANOVA tests to evaluate unadjusted associations between short/long sleep and WMH, FA, and MD. Next, we fitted multivariable logistic and linear regression models to test for associations between sleep duration and WMH presence, WMH volume, FA, and MD, adjusting for relevant covariates. We constructed three adjusted models: Model 1 included age, sex, and race; Model 2 added cardiovascular risk factors to Model 1 (Table 2); and Model 3 incorporated Model 2 variables plus myocardial infarction. When dealing with missing values, we used a complete case approach for all analyses. Secondary analyses evaluated FA and MD in each of the 48 neuroanatomical regions and interaction analyses focused on socio-economic deprivation using the Townsend Deprivation Index.

## Data Availability

UK Biobank data are available through a procedure described at http://www.ukbiobank.ac.uk/using-the-resource/. Data were accessed using project application number 58743.

## Results

Supplementary Figure 1 describes the assembly of the analytic sample. Of the 502,480 UKB participants, 41,434 agreed to participate in a dedicated brain MRI neuroimaging study and had WMH data. After exclusions, the final analytical sample included 39,502 participants (Table 1). The mean time from enrollment to the neuroimaging assessment was 8.9 (SE 1.8) years.

**Table 1.** Baseline characteristics of the studied population.

| Variable | Optimal Sleep<br>N=28,712 | Short Sleep<br>N=8,422 | Long Sleep<br>N=2,368 | P-value |
| --- | --- | --- | --- | --- |
| Age, mean (SD) | 55 (7.6) | 55 (7.2) | 56.50 (7.8) | <0.001 |
| Male, sex (%) | 13,388 (46.6) | 4,153 (49) | 1,018 (42.6) | <0.001 |
| Race, n (%) |  |  |  | <0.001 |
| Black | 128 (0.4) | 124 (1.5) | 6 (0.3) |  |
| Asian | 339 (1.2) | 142 (1.7) | 33 (1.4) |  |
| White | 27,939 (97.5) | 8,053 (95.3) | 2,317 (97.1) |  |
| Other | 243 (0.8) | 127 (1.5) | 31 (1.3) |  |
| Smoking Status, n (%) |  |  |  | <0.001 |
| Never | 17,711 (61.8) | 5,043 (59.5) | 1430 (60) |  |
| Previous | 9,285 (32.4) | 2,821 (33.3) | 809 (34) |  |
| Current | 1,664 (5.8) | 606 (7.2) | 151 (6.3) |  |
| Body mass index, mean (SD) | 26.33 (4.09) | 27.02 (4.42) | 27 (4.44) | <0.001 |
| Hyperlipidemia, n (%) | 2,977 (10.4) | 891 (10.5) | 268 (11.2) | 0.4 |
| Diabetes Mellitus, n (%) | 1,433 (5) | 584 (7) | 190 (8) | <0.001 |
| Hypertension, n (%) | 4,913 (17.1) | 1,656 (19.6) | 522 (22) | <0.001 |
| Myocardial Infarction, n (%) | 764 (2.7) | 205 (2.4) | 58 (2.4) | <0.001 |
| Townsend Deprivation Index (SD) | -1.95 (2.69) | -1.66 (2.82) | -1.86 (2.8) | <0.001 |

**Figure 1:**
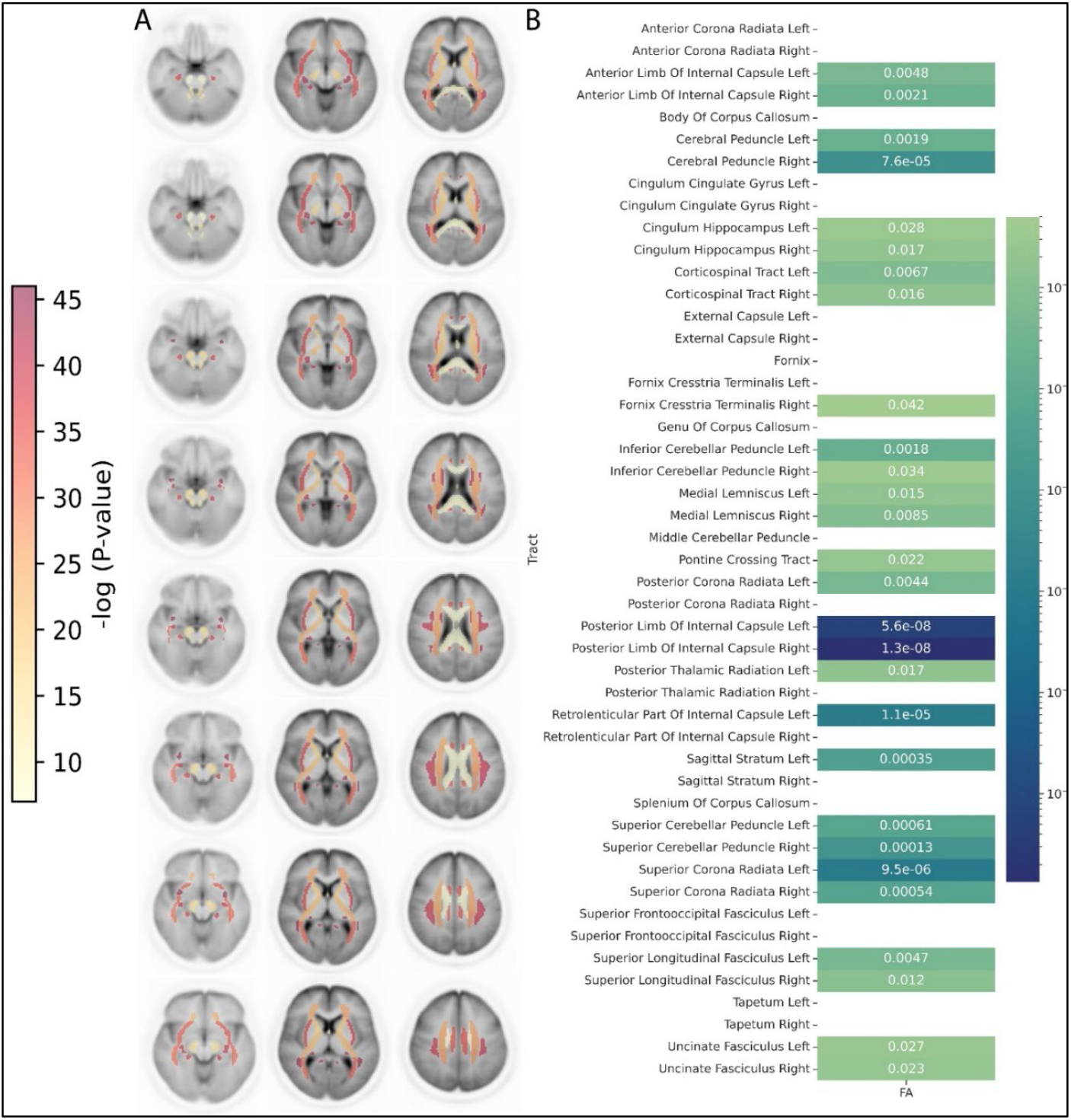
Visual representation of 48 discrete neuroanatomical regions. On the left (A), results from linear regression analysis between short sleep and fractional anisotropy across 48 white matter tracts. On the right (B), a heatmap displays the 48 regions with associated P-Values < 0.05 for the same FA values after regression analysis.

### White Matter Hyperintensities

In unadjusted analyses, the prevalence of WMH was 28.6%, 30.3% and 33.4% for optimal, short, and long sleep, respectively (*P*<0.001, Supplementary Table 1). In multivariable logistic and linear regression analyses, compared to optimal sleep, short sleep was associated with a higher likelihood of WMH presence (Model 1, OR 1.11; 95% CI=1.05-1.17; *P*<0.001) and higher WMH volumes (Model 1, beta 0.06; SE=0.01; *P*<0.001), whereas long sleep was associated with higher WMH volumes (Model 1, beta 0.04; SE=0.02; *P*=0.02) but not with WMH presence (Table 2). When using different multivariable models, results were similar for short sleep, but non-significant for long sleep (Models 2 and 3, Table 2).

**Table 2.** Adjusted logistic and linear regression results for short and long sleep, compared to optimal sleep.

| Model | Sleep duration | White matter hyperintensities presence |  | White matter hyperintensities volume* |  | Fractional anisotropy score |  | Mean diffusivity score |  |
| --- | --- | --- | --- | --- | --- | --- | --- | --- | --- |
|  |  | OR (95%CI) | p-value | Beta (SD) | p-value | Beta (SD) | p-value | Beta (SD) | p-value |
| **Model 1<sup>a</sup>** | Optimal | Reference |  | Reference |  | Reference |  | Reference |  |
|  | Short | 1.11 (1.05-1.17) | <0.001 | 0.06 (0.01) | <0.001 | 0.03 (0.01) | 0.006 | -0.008 (0.01) | 0.4 |
|  | Long | 1.03 (0.93-1.12) | 0.6 | 0.04 (0.02) | 0.02 | 0.07 (0.02) | <0.001 | 0.5 (0.02) | 0.003 |
| **Model 2<sup>b</sup>** | Optimal | Reference |  | Reference |  | Reference |  | Reference |  |
|  | Short | 1.07 (1.01-1.12) | 0.03 | 0.04 (0.01) | <0.001 | 0.02 (0.01) | 0.1 | -0.01 (0.01) | 0.1 |
|  | Long | 0.97 (0.87-1.07) | 0.6 | 0.02 (0.02) | 0.3 | 0.05 (0.01) | 0.005 | 0.04 (0.02) | 0.02 |
| **Model 3<sup>c</sup>** | Optimal sleep | Reference |  | Reference |  | Reference |  | Reference |  |
|  | Short | 1.07 (1.01-1.12) | 0.03 | 0.04 (0.01) | <0.001 | 0.02 (0.01) | 0.2 | -0.02 (0.01) | 0.09 |
|  | Long | 0.97 (0.87-1.07) | 0.6 | 0.02 (0.02) | 0.3 | 0.05 (0.02) | 0.005 | 0.04 (0.02) | 0.02 |

<sup>a</sup> Model 1: covariates = age, sex and race.
<sup>b</sup> Model 2: covariates = model 1+hypertension, diabetes mellitus, smoking status, hyperlipidemia, and body mass index.
<sup>c</sup> Model 3: covariates = model 2+myocardial infarction. \* = natural log transformed.

### Fractional Anisotropy and Mean Diffusivity

In unadjusted analyses, the mean FA scores were -0.02, 0.02 and 0.15 for optimal, short, and long sleep, respectively (*P*<0.001, Supplementary Table 1). In multivariable linear regression analyses, compared to optimal sleep, short (Model 1, *beta*=0.03; *SE*=0.01; *P*=0.006) and long sleep (*beta*=0.07; *SE*=0.02; *P*<0.001) were both associated with higher FA scores. When applying different multivariable models, results were similar for long sleep, but non-significant for short sleep (Models 2 and 3, Table 2). We also found evidence of an association between long sleep and MD scores. In unadjusted analyses, MD scores were -0.01, -0.004 and 0.14 for optimal, short, and long sleep, respectively (*P*<0.001, Supplementary Table 1). In multivariable linear regression analyses, compared to optimal sleep, long sleep (Model 1, *beta* 0.05; *SE*=0.02; *P*=0.003) but not short sleep (Model 1, *beta*=-0.008; *SE*=0.01; *P*=0.4) was associated with the MD score. Similar results were obtained when using different multivariable models (Models 2 and 3, Table 2).

### Additional Analyses

Analyses of the 48 white matter tracts showed associations between short sleep and worse FA profiles in the posterior limbs of the internal capsule (P<0.001, Fig. 1) and long sleep with neuroanatomical FA profile changes in the fornix (P<0.001, Supplementary Fig. 2). No associations were observed between short sleep and MD (Supplementary Fig. 3), but long sleep was associated with worse MD profiles in the genu of the corpus callosum (P<0.001, Supplementary Fig. 4). A significant interaction between long sleep and social disadvantage on FA score was found (interaction P=0.02, data not shown).

## Discussion

We analyzed data from a large neuroimaging study nested within the UK Biobank and found that suboptimal sleep duration, was associated with worse neuroimaging brain health profiles in middle-aged adults without clinically evident neurological disease. Of note, the association between poor sleep and poorer neuroimaging profiles was stronger in persons living in deprived areas.

Mounting evidence points to an important link between sleep duration and cardiovascular health.^1^ Our results extend these prior findings by showing a robust relationship between suboptimal sleep duration and worse brain health, as evaluated via MRI markers. These results are aligned with several converging lines of evidence that indicate that cardiovascular and brain health have significant shared pathophysiology and risk factors.^3^ Because our study included middle-aged persons who have not yet sustained a significant neurological event (stroke or dementia), our results support the addition of sleep to the growing number of cardiometabolic and lifestyle factors that should be optimized during middle-age to improve brain health trajectories later in life. Along these lines, the American Heart Association has recently incorporated sleep duration to the Life’s Simple 7, leading to Life’s Essential 8, a clinical and public health tool developed to highlight the factors that should be corrected to improve cardiovascular and brain health.^2^

In conclusion, our study found that suboptimal sleep duration is associated with silent brain injury in middle-aged adults without a history of neurological disease. Maintaining optimal sleep habits may improve brain health and encourage further research on whether sleep improvement impacts clinical brain health outcomes (e.g., stroke or dementia).

## Supporting information

Full supplemental material (table and figures).

## Data Availability

All data produced are available online at http://www.ukbiobank.ac.uk/using-the-resource/.

## Abbreviations

FA: Fractional Anisotropy
MD: Mean Diffusivity
UKB: United Kingdom Biobank
WMH: White Matter Hyperintensities.

