## Supplementary material for "Suboptimal Sleep Duration is Associated with Poorer Neuroimaging Brain Health Profiles": Full supplemental material (table and figures).

### SUPPLEMENTARY TABLE

**Supplementary Table 1.** Unadjusted results for optimal, short, and long sleep. * = natural log transformed.

| MRI Outcome | Optimal Sleep | Short Sleep | Long Sleep | p-value |
|---|---|---|---|---|
| WMH presence (%) | 8,215 (28.6) | 2,552 (30.3) | 791 (33.4) | <0.001 |
| WMH volume* (SD) | 1.33 (0.99) | 1.40 (0.98) | 1.48 (1.00) | <0.001 |
| FA score (SD) | -0.02 (1.00) | 0.02 (0.99) | 0.15 (1.07) | <0.001 |
| MD score (SD) | -0.01 (1.00) | -0.004 (0.98) | 0.14 (1.07) | <0.001 |

**SUPPLEMENTARY FIGURES:**

**Supplementary Figure 1.**

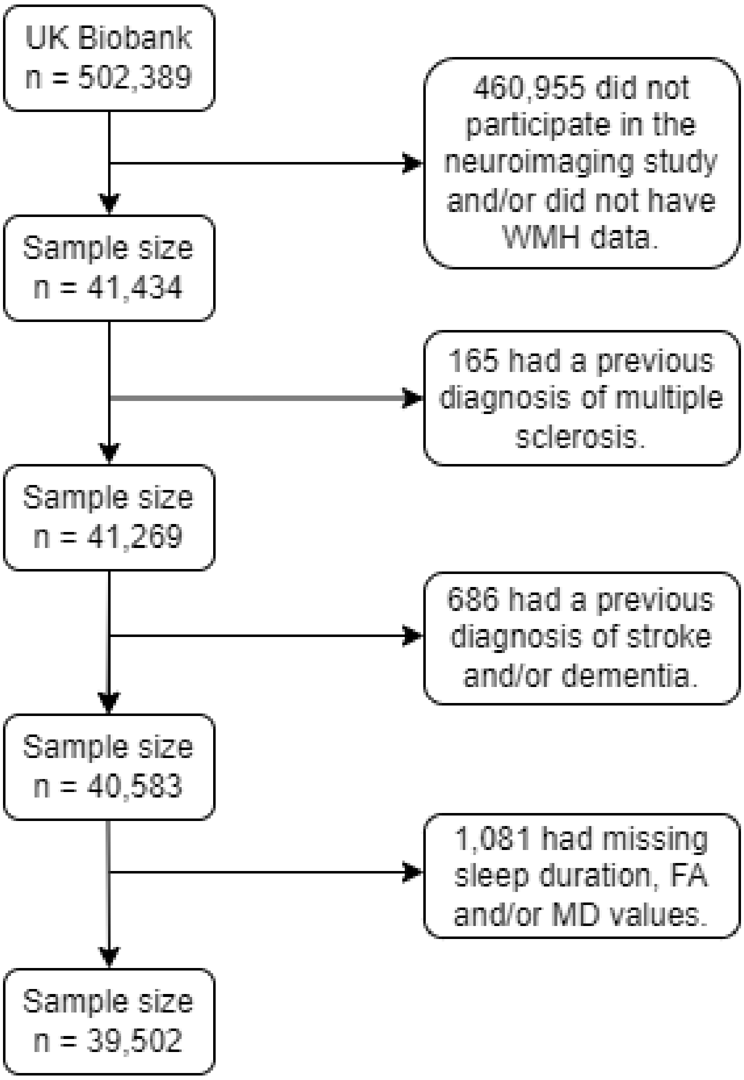


**Supplementary Figure 1: Flowchart summarizing the exclusions that lead to the study population.**

**Supplementary Figure 2.**

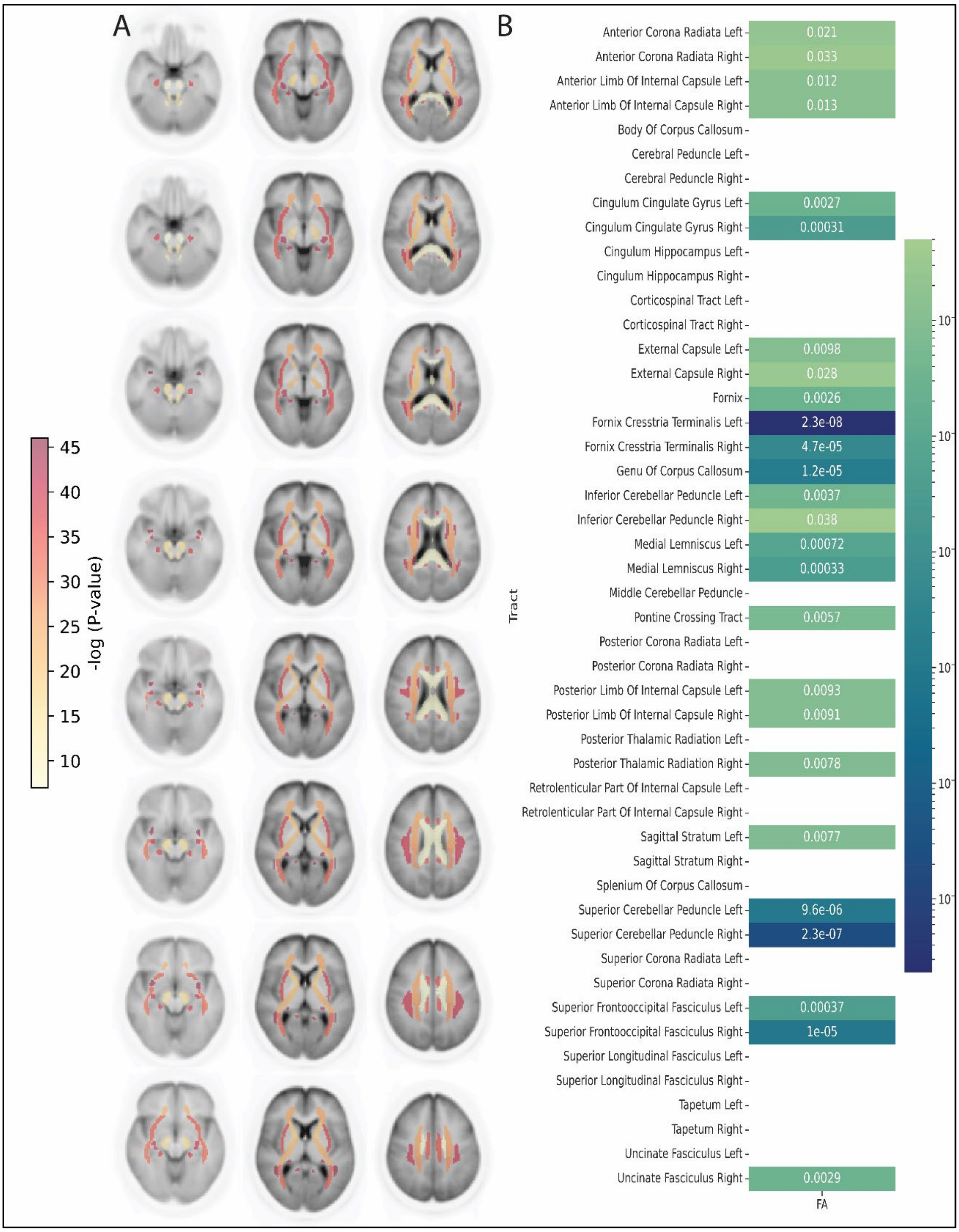


**Supplementary Figure 2: Visual representation of 48 discrete neuroanatomical regions.**

On the left, (A), results from linear regression analysis between long sleep and fractional anisotropy across 48 discrete white matter tracts. On the right, (B), a heatmap portraying the 48 neuroanatomical regions and their associated P-Values < 0.05 after the same regression analysis, for the same fractional anisotropy values.

**Supplementary Figure 3.**

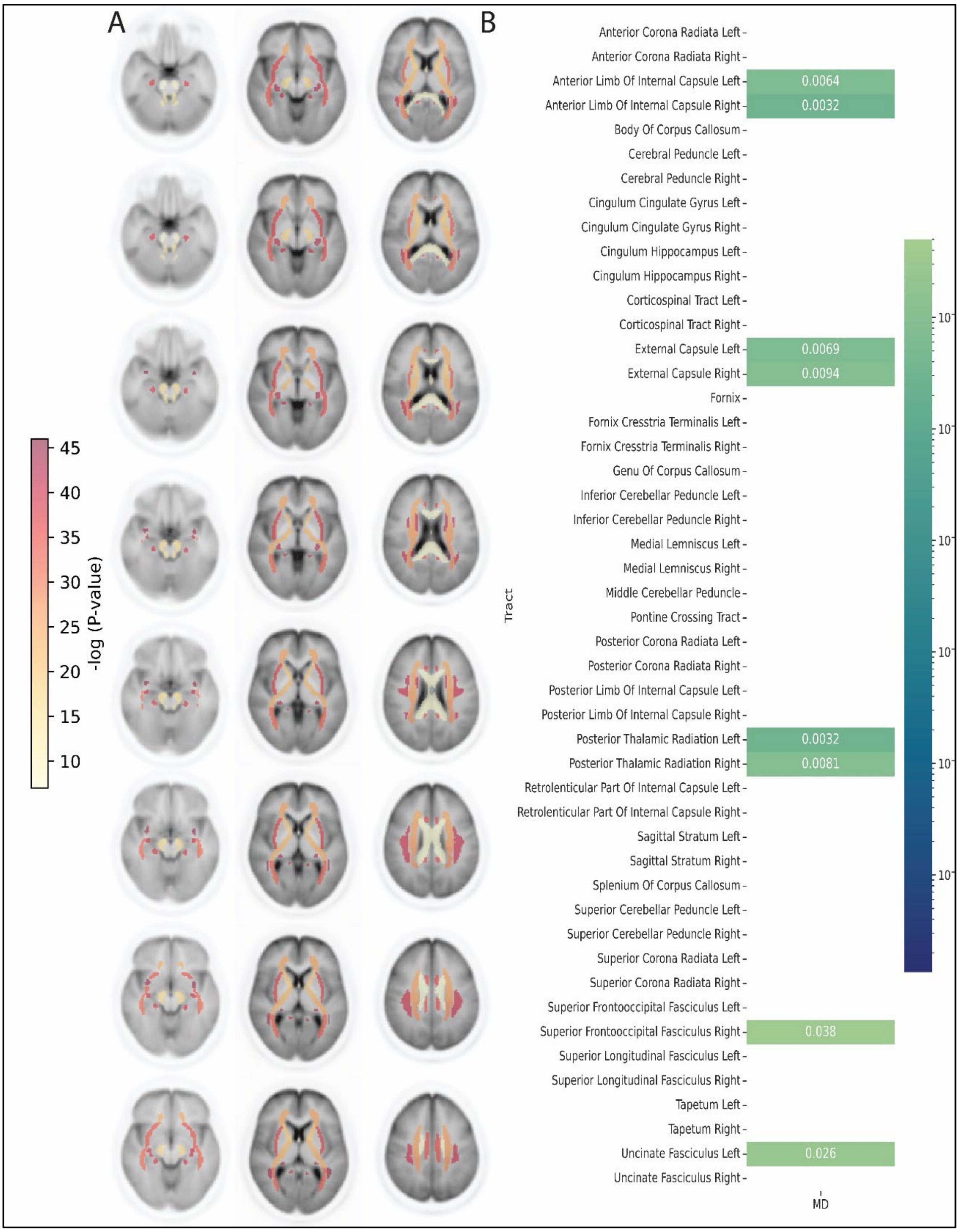


**Supplementary Figure 3: Visual representation of 48 discrete neuroanatomical regions.**

On the left, (A), results from linear regression analysis between short sleep and mean diffusivity across 48 discrete white matter tracts. On the right, (B), a heatmap portraying the 48 neuroanatomical regions and their associated P-Values < 0.05 after the same regression analysis, for the same fractional anisotropy values.

**Supplementary Figure 4.**

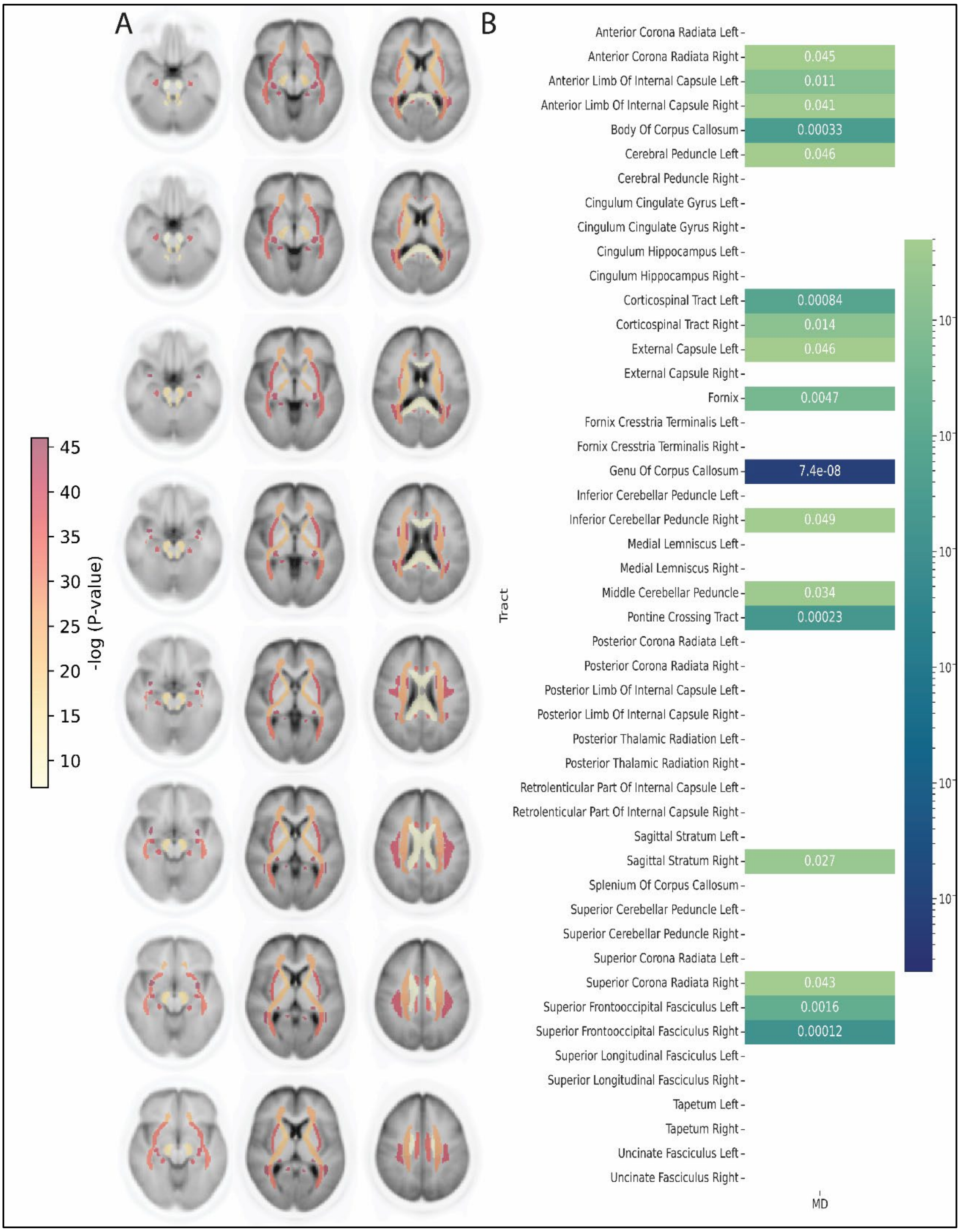


**Supplementary Figure 4: Visual representation of 48 discrete neuroanatomical regions.**

On the left, (A), results from linear regression analysis between long sleep and mean diffusivity across 48 discrete white matter tracts. On the right, (B), a heatmap portraying the 48 neuroanatomical regions and their associated P-Values < 0.05 after the same regression analysis, for the same fractional anisotropy values.
